# Navigating Visual Challenges: How Parkinson’s Disease Alters Cognitive Priorities in Visual Search

**DOI:** 10.1101/2024.02.19.24303067

**Authors:** Sinem Balta Beylergil, Peggy Skelly, Ibrahim Quagraine, Camilla Kilbane, Fatema F. Ghasia, Aasef G. Shaikh

## Abstract

**Objective:** Parkinson’s disease (PD) hampers visual control in tasks such as reading, driving, and navigation. This study explores the impact of contextual visual priors on visual search behavior in PD, examining expectations from past experiences and guiding cognitive processes.

**Methods:** We compared eye movements during a visual search task with complex scenes to evaluate gaze strategies in PD, compared to healthy controls.

**Results:** PD participants prolonged fixation on high-probability areas for the target object, consistent across expected and unexpected locations. The emphasis on visual priors was proven beneficial in expected locations but presented challenges when the target was situated in an unlikely place.

**Conclusion:** These findings indicate that PD alters attention allocation and visual processing by affecting the utilization of contextual visual priors. This study contributes to understanding how PD impacts visual search behavior and cognitive processing, providing insights for potential interventions targeting visual deficits in PD patients.

## Introduction

People with Parkinson’s disease (PD) frequently report problems with navigational visual tasks, such as driving or reading^1, 2^. The deficits stem from many causes. One of the causes manifests as an impairment in higher visual function, such as the interplay of top-down process such as visual reaction time and adaptation of prior knowledge based on contextual cues (contextual visual priors)^3–8^. The classic example is impaired performance of modified Tower of London task that requires the utilization of the bottom up visual information, forming working memory, and devising effective solutions to overcome obstacles by adjusting the initial strategy^9^.

Contextual prior information, acting as a top-down influence, encompasses an individual’s knowledge and expectations in visual tasks. Derived from past experiences, it guides and biases visual perception, shaping the interpretation and processing of sensory information. We hypothesize that PD affects utilization of contextual visual priors, the utilization is abnormally stronger compared to healthy controls.

## Methods

### Subjects

We studied 13 PD men and seven age-matched healthy controls (HC) (PD: mean age 67.5±9.2 years, disease duration 11.5±5.1 years; HC: 4 men, 3 women, mean age 64.3±7.5, p=0.46, unpaired t-test, **Table 1**). PD inclusion criteria were Hoehn and Yahr stage 2–4 when off medication, stable therapeutic regimen, normal corrected visual acuity, and color vision. Exclusion criteria were dementia, untreated psychiatric conditions, structural brain lesions, or atypical parkinsonism. Informed consent was obtained, and the protocol was approved by local institutional review boards. Experiments were done in medication on phase.

### Experimental protocol

The experiment involved a visual search task where the participants identified targets by performing a mouse click at their locations. We simultaneously measured position of the right and left eyes. Visual stimuli from the BOiS-Berlin Object in Scene Database ensured unbiased assessment of human visual search behavior.^10^ The task included 16 trials in 4 blocks, with calibrated eye positions. Each trial presented a scene resembling a familiar environment with a relevant target object. Half had the target in the expected location (e.g.,**Fig.1A**), while others had it in an unexpected location (e.g.,**Fig.1B**). Subjects received a 10-second visual cue, followed by a 45-second view of the scene, terminated upon successful identification or feedback for continued searching after an incorrect mouse click.

**Fig 1.**
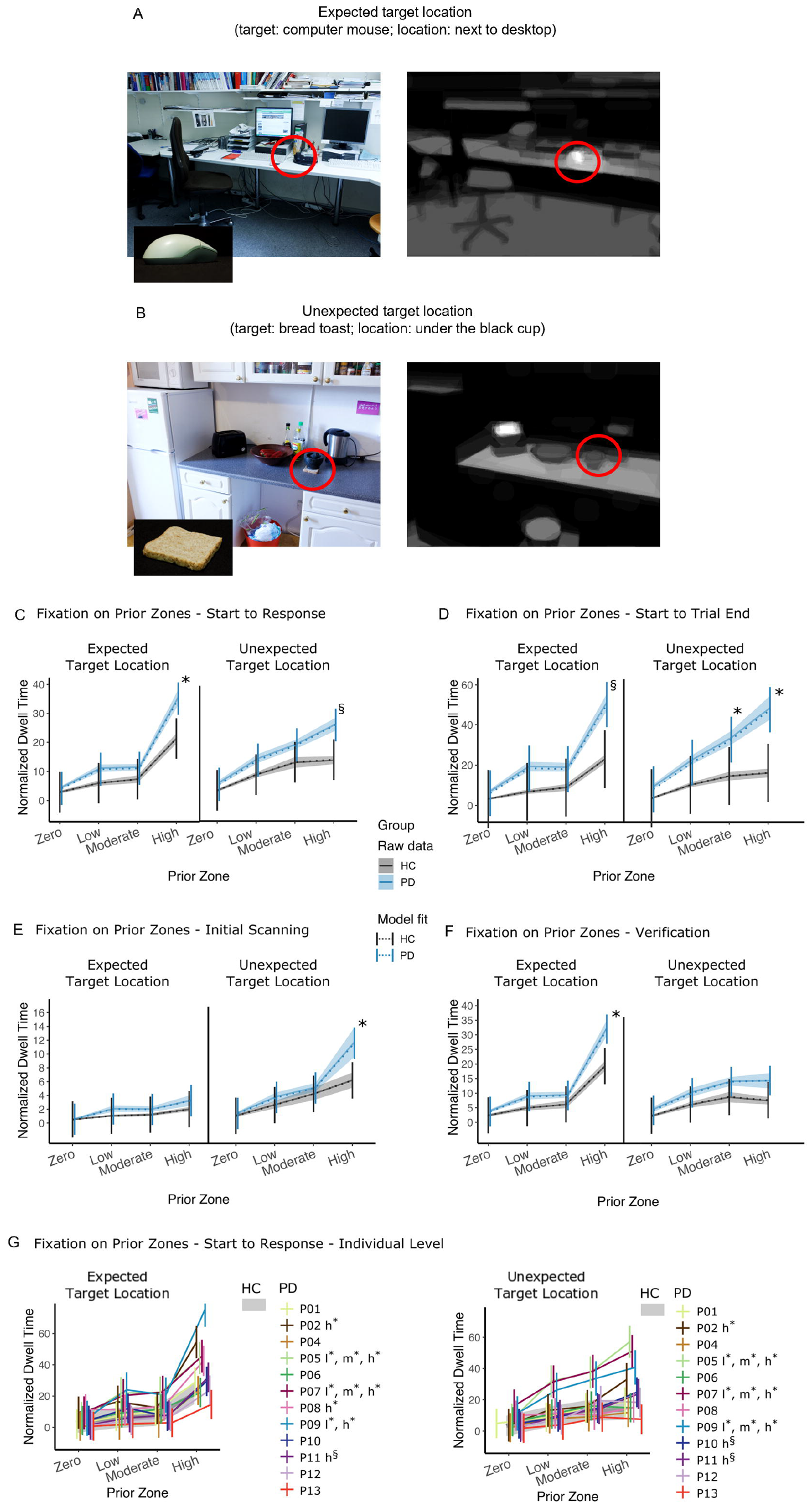
**(A)** An example trial with the target object placed at an expected location. This trial presents an office desk where the target cue (computer mouse) is placed on the desk near the desktop computer. The mouse (circled in red) is at an expected location for this object. This location is an expected location as depicted in bright white signal in the panel on the right. **(B)** An example trial of the task with the target object placed at an unexpected location. This trial presents kitchen counter, where the target cue, slice of bread toast, is located below the black cup on the counter, an unlikely place to find the object. The likely place, is within the toaster oven, as depicted in the right panel with bright white signal. **C-F:** Effect of prior information on fixation at the group level. Between-group comparisons of fixation on zero, low, moderate, and high prior zones of the scenes. Total dwell times per zone normalized by the area percentage of the prior zone (ratio of the zone’s area to the scene’s total area) were compared between HC and PD for the trials with expected target locations (left) and unexpected target locations (right) for four different time windows: **C**. trial start to first response, **D**. trial start to trial end which was determined by either a correct response or automatic trial elapse at 45 s, **E**. during initial scanning, defined as the period between the trial start and the time of the first look on target ROI, and **F**. during verification, defined as the period between the first look on target ROI to the first response. Solid lines with shaded areas display raw data (with standard error) whereas dotted lines are the LMM-predictions with error bars depicting 95% confidence intervals of the predictions. **G**. Effect of Prior Information on Individual Fixation. PD to reference HC comparisons of fixation on zero, low, moderate, and high prior zones of the scenes from the start to the time of first response. LMM predictions of PD subjects are plotted with the mean HC predictions (thick black lines, shaded area: 95% CI) for trials with the target object placed at an expected location (left) or an unexpected location (right). Letters l (low), m (moderate), and h (high) with asterisks next to the subject IDs indicate a significant difference between the subject compared to the reference control subjects for the low, moderate, and high prior zones, respectively. * indicates p<.05 Holm-corrected for multiple comparisons. § indicates p<.01 uncorrected for multiple comparisons.

### Data Analysis

Trial accuracy was determined by a 100-pixel circular ROI around the target object, measuring response time (RT) from scene presentation to ROI click. A mixed ANOVA compared PD and HC considering group, target location, and viewing condition. Linear mixed-effects models (LMM) analyzed RT, including group, target location, interaction, viewing condition, subject ID, and trial index. LMM compared PD to HC with post hoc Holm-corrected analyses. Type III ANOVA p-values examined group differences in initial ROI glance (t_initial_) and ROI glance to RT (RT-t_initial_). Spearman’s correlations assessed RT, t_initial_, RT-t_initial_, and subject scores. Rstatix, stats, and ggplot packages in R were used for analysis and visualization.

### Fixation on Target Object

Fixation duration on the target object within a 100-pixel radius circular ROI (ROI dwell time) was analyzed using concise LMMs: Dwell Time ~ Group * Target Location + Viewing + Saliency + (1|Trial ID) + (1|Subject ID). Excluding saccadic eye movement intervals, data were analyzed from trial start to the first response (RT) or trial conclusion. Viewing condition and average ROI saliency were covariates.

### Fixation Maps and Contextual Prior Map Analysis

The contextual prior maps, derived from the BOiS Database, quantitatively captured common expectations for locating a target object (for details, Mohr et al., 2016). For example grayscale maps in **Fig.1A,B** depict likelihoods of target object appearance (white: most-likely; black: least-likely). Scenes were divided into zones based on pixel values (zero, low, moderate, high), with resulting Boolean masks calculated total fixation duration (**Supplementary Fig.S1**).

Normalized dwell times were analyzed using LMM, considering group (PD and HC), target location, and prior zone factors with their three-way interaction. Covariates included viewing condition and average saliency of each prior zone. Random effects incorporated subject ID and trial index (Normalized Dwell Time ~ Group * Prior Zone * Target Location + Viewing + Saliency + (1|Trial ID) + (1|Subject ID)). Analysis covered RT and trial initiation to trial end. Group differences in normalized dwell times were assessed in two intervals: initial scanning (trial start to first glance at target ROI) and verification scanning (first ROI glance to RT). Individual-level analyses used LMM for ROI and prior zone dwell times in 12 of 13 PD patients, comparing each patient’s fixation durations to the HC group, with a consistent model framework, one remaining patient had only first four trials.

## Results

### Exploring Visual Search: Group-level Study on Fixations in Prior Zones

PD influenced the utilization of contextual prior information in goal-directed visual search. Analyzing dwell time from trial start to the first response revealed significant group by prior zone effect (F(3, 2254.31)=11.783, p<0.001). PD showed longer fixation duration on high prior zones for all trials (F(1,19.35)=4.914, p=0.039) in expected as well as unexpected locations (**Fig.1C**). Excluding target ROI pixels also indicated longer fixations in PD for high prior zones (group by prior zone interaction: F(3,2254.08)=12.901, p<0.001; group effect in high prior zone: F(1,19.46)=5.130, p=0.035). Zero, low, and moderate prior zones showed no significant difference between groups (p>0.05).

In the dwell time analysis from trial start to end, PD exhibited higher fixation durations on areas with more prior information (group by prior zone interaction:F(3, 2254.73)=16.699, p<0.001) (**Fig.1D**). PD showed longer fixations on high and moderate prior zones (F(1, 19.65)=4.901, p=0.039 and F(1,549.11)=4.303, p=0.039), particularly for trials with unexpected targets (moderate: p=0.047, high: p=0.041, Holm-corrected), while zero and low prior zones showed no significant difference (p>0.05).

During the initial scanning period (trial start to first look on target ROI), a significant group by prior zone interaction was found (F(3, 2257.49)= 2.856, p=0.036; **Fig.1E**). Pairwise comparisons revealed higher fixations on high prior zones in PD during unexpected target trials (p=0.024, Holm-corrected), with no significant group differences in other prior zones (p>0.05).

In the verification scanning period (first look on target ROI to first response), PD exhibited longer fixations on high prior zones only for expected target trials (p=0.028), as shown by a significant group by prior zone interaction (F(3,2255.28)=5.162, p=0.001; **Fig.1F**).

### Visual Search: Correlation Between Fixations on High Prior Zones and Scores

Mean fixation durations on high prior zones (normalized by total duration) were positively correlated with mean scores in expected target location trials (r(32)=0.442, p=0.011) and negatively correlated with mean scores in unexpected target location trials (r(32)=-.542, p=0.001) across the entire group. The longer initial scanning period in PD during trials with unexpected targets was negatively correlated with scores (r(13)=-0.762, p=0.002, **Fig.1E**). Similarly, PD’s longer fixations on high prior zones during the verification scanning period in trials with expected targets were positively correlated with mean scores in those trials (r(13)=0.593, p=0.033, **Fig.1F**). *Visual Search: Analyzing Individual Fixations in Prior Zones:*

Individual fixational data of 12 PD patients were analyzed using independent LMMs, comparing them to reference HC groups (excluding one participant with incomplete data). Normalized dwell times, from trial start to the first response, were the response variable. Fixed-effect factors included group, prior zone, and target location, with random-effect factors of subject ID and trial ID. Covariates were viewing condition and mean saliency. Seven patients showed significant group or group by prior zone effects (**Supplementary Table S1**). Four had longer fixations on high prior zones for both target locations (p<.05, Holm-corrected, **Fig.1G**). One patient exhibited longer dwell times only for expected target locations. Two patients showed heightened dwell times for unexpected targets, and one for both expected and unexpected target locations. Additionally, three patients demonstrated prolonged dwell times for low and moderate prior zones (p<.05, Holm-corrected; **Fig.1G**).

## Discussion

This study explores how contextual prior information affects visual search behavior in PD. Prolonged fixations on high prior zones in PD suggest altered processing of contextual cues, indicating potential shifts in attention distribution and visual processing. Consistent extended fixations across target locations and search phases imply a general alteration in how PD patients interact with their visual environment, reflecting changes in perceptual processing. Correlation analysis reveals that fixation on high prior zones correlates with target identification scores. Longer fixations are linked to higher scores in trials with expected targets but lower scores in trials with unexpected targets, suggesting unproductive attention allocation based on prior knowledge. The negative correlation with extended initial scanning in unexpected target trials highlights its detrimental impact. Results suggest challenges for PD patients in adapting to unexpected scenarios, aligning with “cognitive and perceptual rigidity.” Positive correlation between longer fixations during verification scanning and scores in trials with expected targets indicates a beneficial role. This nuanced relationship emphasizes that fixational behavior effects on task performance are influenced by target location and scanning phase.

The altered contextual visual priors in PD suggest that visual perception results from the interplay between external sensory input (bottom-up) and internal representations scenes (top-down)^11–15^. The dominance of bottom-up or top-down bias the information determining which proto-object enters conscious awareness. Bottom-up bias is dominant for objects with distinct physical properties, while top-down bias is influenced by familiarity, goals, intentions, and spatial attention – visual “priors” based on long-term memory of scene perceptions^16^. PD induces a top-down bias by impacting the lateral frontal cortex and ventral visual stream. The thalamus, crucial for relaying information to the occipital cortex and ventral visual stream, plays a key role^17, 18^.

Cholinergic cells in the nucleus basalis of Meynert project to the frontal cortex, ventral visual stream, and thalamic nuclei^19, 20^. Brainstem cholinergic projections from pedunculo-pontine nucleus and laterodorsal tegmental nucleus modulate various regions, including the reticular formation, mediodorsal nucleus, lateral geniculate, and occipital cortex^21^. The connectivity pattern among basal ganglia, pedunculo-pontine nucleus, and nucleus basalis of Meynert influences the lateral frontal cortex and ventral visual stream, forming the basis for altered contextual visual priors in PD. The study tested the hypothesis that PD enhances utilization of contextual visual priors, showing their increased strength in the disease state compared to controls.

## Conclusion

The study provides valuable insights into how PD influences visual search behavior, highlighting changes in fixational behavior and their impact on task performance. The findings contribute to understanding cognitive and perceptual changes in PD, with implications for designing interventions to address visual processing challenges in individuals with PD.

## Data Availability

Data will be available to interested party after appropriate institutional formalities are completed between requesting institution and the US Department of Veterans Affairs.

## Acknowledgments

The authors thank Dr. Jonathan Jacobs and Dr.Jordan Murray for collegial support.

## Authors’ roles

A.Shaikh, S.B.Beylergil, and F. Ghasia conceptualized the study and wrote the manuscript. All authors collected data, analyzed data, and edited manuscript.

## Figure legends

**Supplemental Figure 1**. An example trial with a watering can as the target object (target), the planter as the unexpected target location, and the room as the scene (original scene). The gray scale contextual prior map quantifies the likelihood of the pixels of the scene to contain the target object with a scale varying between 0 (black, zero likelihood) and 255 (white, highest likelihood). Four black and white Boolean maps on the right were derived from the contextual prior map (see the arrows) by segmenting the pixels based on their values. Black pixels with value 0 were included in the zero prior zone, the remaining pixel values were split into three equal sizes using quartiles (33.33% and 66.66%) forming the low, moderate and high prior zones. Resulting maps were used to calculate the fixation duration of each subject on the four prior zones for each trial after normalizing the duration by the total number of black pixels in zones.

