## Supplementary material for "Navigating Visual Challenges: How Parkinson’s Disease Alters Cognitive Priorities in Visual Search": Table s1

**Table 3. PD vs. HC comparisons of fixation on prior zones at individual level.**

| **P01** |  |  |  |  |
| --- | --- | --- | --- | --- |
|  | **Num df** | **Den df** | **F value** | **p** |
| **Target location** | 1 | 46.646 | 0.000 | 0.983 |
| **Group** | 1 | 7.924 | 0.334 | 0.580 |
| **PriorZone** | 3 | 972.827 | 46.570 | **<0.001** |
| **Viewing** | 1 | 31.457 | 2.985 | 0.094 |
| **Target location:Group** | 1 | 973.006 | 0.089 | 0.765 |
| **Target location:PriorZone** | 3 | 972.827 | 8.021 | **<0.001** |
| **Group:PriorZone** | 3 | 972.827 | 1.578 | 0.193 |
| **Target location:Group:PriorZone** | 3 | 972.827 | 0.237 | 0.871 |
| **Target Location = expected** | | | |  |
|  | **Chi-square** | **Df** | **p** |  |
| **Group** | 0.464 | 1 | 0.496 |  |
| **PriorZone** | 200.674 | 3 | **<0.001** |  |
| **Viewing** | 0.166 | 1 | 0.684 |  |
| **Group:PriorZone** | 3.808 | 3 | 0.283 |  |
| **Target location = unexpected** | | | |  |
|  | **Chi-square** | **Df** | **p** |  |
| **Group** | 0.207 | 1 | 0.649 |  |
| **PriorZone** | 76.043 | 3 | **<0.001** |  |
| **Viewing** | 4.099 | 1 | **0.043** |  |
| **Group:PriorZone** | 1.439 | 3 | 0.696 |  |
| **Pairwise comparisons (Target location = expected)** | | | | |
| **contrast** | **df** | **t value** | **uncorrected p** | **Holm-corrected p** |
| **HC zero - PD zero** | 21.163 | 0.151 | 0.881 | 1.000 |
| **HC low - PD low** | 21.163 | -0.192 | 0.850 | 1.000 |
| **HC moderate - PD moderate** | 21.163 | -0.580 | 0.568 | 1.000 |
| **HC high - PD high** | 21.163 | -1.433 | 0.166 | 0.665 |
| **Pairwise comparisons (Target location = unexpected)** | | | | |
| **contrast** | **df** | **t value** | **uncorrected p** | **Holm-corrected p** |
| **HC zero - PD zero** | 16.623 | -0.266 | 0.793 | 1.000 |
| **HC low - PD low** | 16.623 | -0.011 | 0.991 | 1.000 |
| **HC moderate - PD moderate** | 16.623 | -0.313 | 0.758 | 1.000 |
| **HC high - PD high** | 16.623 | -0.853 | 0.406 | 1.000 |
| **P02** |  |  |  |  |
|  | **Num df** | **Den df** | **F value** | **p** |
| **Target location** | 1 | 56.701 | 2.281 | 0.137 |
| **Group** | 1 | 8.183 | 6.152 | **0.037** |
| **PriorZone** | 3 | 952.804 | 66.421 | **<0.001** |
| **Viewing** | 1 | 31.538 | 2.085 | 0.159 |
| **Target location:Group** | 1 | 960.755 | 6.795 | **0.009** |
| **Target location:PriorZone** | 3 | 952.804 | 8.820 | **<0.001** |
| **Group:PriorZone** | 3 | 952.804 | 15.643 | **<0.001** |
| **Target location:Group:PriorZone** | 3 | 952.804 | 0.803 | 0.492 |
| **Target location = expected** | | | |  |
|  | **Chi-square** | **Df** | **p** |  |
| **Group** | 11.720 | 1 | **0.001** |  |
| **PriorZone** | 164.453 | 3 | **<0.001** |  |
| **Viewing** | 0.144 | 1 | 0.704 |  |
| **Group:PriorZone** | 30.513 | 3 | **<0.001** |  |
| **Target location = unexpected** | | | |  |
|  | **Chi-square** | **Df** | **p** |  |
| **Group** | 2.153 | 1 | 0.142 |  |
| **PriorZone** | 72.857 | 3 | **<0.001** |  |
| **Viewing** | 2.877 | 1 | 0.090 |  |
| **Group:PriorZone** | 17.231 | 3 | **<0.001** |  |
| **Pairwise comparisons (Target location = expected)** | | | | |
| **contrast** | **df** | **t value** | **uncorrected p** | **Holm-corrected p** |
| **HC zero - PD zero** | 30.710 | -1.134 | 0.266 | 0.531 |
| **HC low - PD low** | 30.710 | -1.807 | 0.081 | 0.242 |
| **HC moderate - PD moderate** | 30.710 | -0.898 | 0.376 | 0.531 |
| **HC high - PD high** | 30.710 | -5.761 | **<0.001** | **<0.001** |
| **Pairwise comparisons (Target location = unexpected)** | | | | |
| **contrast** | **df** | **t value** | **uncorrected p** | **Holm-corrected p** |
| **HC zero - PD zero** | 20.684 | -0.070 | 0.945 | 1.000 |
| **HC low - PD low** | 20.684 | -0.743 | 0.466 | 1.000 |
| **HC moderate - PD moderate** | 20.684 | -0.474 | 0.641 | 1.000 |
| **HC high - PD high** | 20.684 | -3.197 | **0.004** | **0.018** |
| **P04** |  |  |  |  |
|  | **Num df** | **Den df** | **F value** | **p** |
| **Target location** | 1 | 45.228 | 0.077 | 0.782 |
| **Group** | 1 | 7.923 | 0.274 | 0.615 |
| **PriorZone** | 3 | 972.747 | 39.882 | **<0.001** |
| **Viewing** | 1 | 31.389 | 2.100 | 0.157 |
| **Target location:Group** | 1 | 972.913 | 0.017 | 0.895 |
| **Target location:PriorZone** | 3 | 972.747 | 11.655 | **<0.001** |
| **Group:PriorZone** | 3 | 972.747 | 0.830 | 0.477 |
| **Target location:Group:PriorZone** | 3 | 972.747 | 0.613 | 0.607 |
| **Target location = expected** | | | |  |
|  | **Chi-square** | **Df** | **p** |  |
| **Group** | 0.338 | 1 | 0.561 |  |
| **PriorZone** | 222.504 | 3 | **<0.001** |  |
| **Viewing** | 0.029 | 1 | 0.866 |  |
| **Group:PriorZone** | 3.547 | 3 | 0.315 |  |
| **Target location = unexpected** | | | |  |
|  | **Chi-square** | **Df** | **p** |  |
| **Group** | 0.195 | 1 | 0.659 |  |
| **PriorZone** | 70.617 | 3 | **<0.001** |  |
| **Viewing** | 3.178 | 1 | 0.075 |  |
| **Group:PriorZone** | 0.676 | 3 | 0.879 |  |
| **Pairwise comparisons (Target location = expected)** | | | | |
| **contrast** | **df** | **t value** | **uncorrected p** | **Holm-corrected p** |
| **HC zero - PD zero** | 19.795 | 0.442 | 0.663 | 1.000 |
| **HC low - PD low** | 19.795 | 0.872 | 0.393 | 1.000 |
| **HC moderate - PD moderate** | 19.795 | 0.908 | 0.375 | 1.000 |
| **HC high - PD high** | 19.795 | -0.444 | 0.662 | 1.000 |
| **Pairwise comparisons (Target location = unexpected)** | | | | |
| **contrast** | **df** | **t value** | **uncorrected p** | **Holm-corrected p** |
| **HC zero - PD zero** | 16.197 | 0.218 | 0.830 | 1.000 |
| **HC low - PD low** | 16.197 | 0.164 | 0.872 | 1.000 |
| **HC moderate - PD moderate** | 16.197 | 0.695 | 0.497 | 1.000 |
| **HC high - PD high** | 16.197 | 0.333 | 0.743 | 1.000 |
| **P05** |  |  |  |  |
|  | **Num df** | **Den df** | **F value** | **p** |
| **Target location** | 1 | 49.976 | 5.188 | **0.027** |
| **Group** | 1 | 7.891 | 21.518 | **0.002** |
| **PriorZone** | 3 | 972.822 | 79.100 | **<0.001** |
| **Viewing** | 1 | 31.558 | 2.125 | 0.155 |
| **Target location:Group** | 1 | 973.037 | 12.312 | **<0.001** |
| **Target location:PriorZone** | 3 | 972.822 | 2.530 | 0.056 |
| **Group:PriorZone** | 3 | 972.822 | 20.280 | **<0.001** |
| **Target location:Group:PriorZone** | 3 | 972.822 | 2.303 | 0.076 |
| **Target location = expected** | | | |  |
|  | **Chi-square** | **Df** | **p** |  |
| **Group** | 12.986 | 1 | 0.000 |  |
| **PriorZone** | 165.067 | 3 | 0.000 |  |
| **Viewing** | 0.035 | 1 | 0.851 |  |
| **Group:PriorZone** | 15.001 | 3 | 0.002 |  |
| **Target location = unexpected** | | | |  |
|  | **Chi-square** | **Df** | **p** |  |
| **Group** | 27.637 | 1 | **<0.001** |  |
| **PriorZone** | 76.677 | 3 | **<0.001** |  |
| **Viewing** | 3.181 | 1 | 0.075 |  |
| **Group:PriorZone** | 49.917 | 3 | **<0.001** |  |
| **Pairwise comparisons (Target location = expected)** | | | | |
| **contrast** | **df** | **t value** | **uncorrected p** | **Holm-corrected p** |
| **HC zero - PD zero** | 25.831 | -0.576 | 0.570 | 0.570 |
| **HC low - PD low** | 25.831 | -2.910 | **0.007** | **0.022** |
| **HC moderate - PD moderate** | 25.831 | -2.723 | **0.011** | **0.023** |
| **HC high - PD high** | 25.831 | -4.194 | **<0.001** | **0.001** |
| **Pairwise comparisons (Target location = unexpected)** | | | | |
| **contrast** | **df** | **t value** | **uncorrected p** | **Holm-corrected p** |
| **HC zero - PD zero** | 23.346 | -0.625 | 0.538 | 0.538 |
| **HC low - PD low** | 23.346 | -3.778 | **0.001** | **0.002** |
| **HC moderate - PD moderate** | 23.346 | -4.012 | **0.001** | **0.002** |
| **HC high - PD high** | 23.346 | -7.069 | **<0.001** | **<0.001** |
| **P06** |  |  |  |  |
|  | **Num df** | **Den df** | **F value** | **p** |
| **Target location** | 1 | 48.482 | 0.016 | 0.900 |
| **Group** | 1 | 7.937 | 0.332 | 0.580 |
| **PriorZone** | 3 | 972.838 | 29.917 | **<0.001** |
| **Viewing** | 1 | 31.420 | 4.486 | **0.042** |
| **Target location:Group** | 1 | 973.034 | 0.013 | 0.908 |
| **Target location:PriorZone** | 3 | 972.838 | 6.311 | **<0.001** |
| **Group:PriorZone** | 3 | 972.838 | 0.275 | 0.844 |
| **Target location:Group:PriorZone** | 3 | 972.838 | 0.074 | 0.974 |
| **Target location = expected** | | | |  |
|  | **Chi-square** | **Df** | **p** |  |
| **Group** | 0.399 | 1 | 0.528 |  |
| **PriorZone** | 187.393 | 3 | **<0.001** |  |
| **Viewing** | 0.507 | 1 | 0.477 |  |
| **Group:PriorZone** | 0.788 | 3 | 0.852 |  |
| **Target location = unexpected** | | | |  |
|  | **Chi-square** | **Df** | **p** |  |
| **Group** | 0.247 | 1 | 0.619 |  |
| **PriorZone** | 68.856 | 3 | **<0.001** |  |
| **Viewing** | 4.785 | 1 | **0.029** |  |
| **Group:PriorZone** | 0.235 | 3 | 0.972 |  |
| **Pairwise comparisons (Target location = expected)** | | | | |
| **contrast** | **df** | **t value** | **uncorrected p** | **Holm-corrected p** |
| **HC zero - PD zero** | 20.998 | -0.275 | 0.786 | 1.000 |
| **HC low - PD low** | 20.998 | -0.630 | 0.536 | 1.000 |
| **HC moderate - PD moderate** | 20.998 | -0.828 | 0.417 | 1.000 |
| **HC high - PD high** | 20.998 | -0.177 | 0.861 | 1.000 |
| **Pairwise comparisons (Target location = unexpected)** | | | | |
| **contrast** | **df** | **t value** | **uncorrected p** | **Holm-corrected p** |
| **HC zero - PD zero** | 16.841 | -0.457 | 0.653 | 1.000 |
| **HC low - PD low** | 16.841 | -0.367 | 0.718 | 1.000 |
| **HC moderate - PD moderate** | 16.841 | -0.544 | 0.594 | 1.000 |
| **HC high - PD high** | 16.841 | -0.205 | 0.840 | 1.000 |
| **P07** |  |  |  |  |
|  | **Num df** | **Den df** | **F value** | **p** |
| **Target location** | 1 | 66.755 | 6.813 | **0.011** |
| **Group** | 1 | 7.942 | 23.428 | **0.001** |
| **PriorZone** | 3 | 972.364 | 49.257 | **<0.001** |
| **Viewing** | 1 | 31.152 | 1.640 | 0.210 |
| **Target location:Group** | 1 | 972.712 | 10.276 | **0.001** |
| **Target location:PriorZone** | 3 | 972.364 | 2.876 | **0.035** |
| **Group:PriorZone** | 3 | 972.364 | 8.657 | **<0.001** |
| **Target location:Group:PriorZone** | 3 | 972.364 | 0.230 | 0.875 |
| **Target location = expected** | | | |  |
|  | **Chi-square** | **Df** | **p** |  |
| **Group** | 14.743 | 1 | **<0.001** |  |
| **PriorZone** | 162.959 | 3 | **<0.001** |  |
| **Viewing** | 0.363 | 1 | 0.547 |  |
| **Group:PriorZone** | 11.671 | 3 | **0.009** |  |
| **Target location = unexpected** | | | |  |
|  | **Chi-square** | **Df** | **p** |  |
| **Group** | 29.608 | 1 | **<0.001** |  |
| **PriorZone** | 44.479 | 3 | **<0.001** |  |
| **Viewing** | 5.310 | 1 | **0.021** |  |
| **Group:PriorZone** | 14.234 | 3 | **0.003** |  |
| **Pairwise comparisons (Target location = expected)** | | | | |
| **contrast** | **df** | **t value** | **uncorrected p** | **Holm-corrected p** |
| **HC zero - PD zero** | 27.503 | -1.172 | 0.251 | 0.251 |
| **HC low - PD low** | 27.503 | -2.609 | **0.015** | **0.030** |
| **HC moderate - PD moderate** | 27.503 | -2.761 | **0.010** | **0.030** |
| **HC high - PD high** | 27.503 | -4.414 | **<0.001** | **0.001** |
| **Pairwise comparisons (Target location = unexpected)** | | | | |
| **contrast** | **df** | **t value** | **uncorrected p** | **Holm-corrected p** |
| **HC zero - PD zero** | 31.482 | -1.953 | 0.060 | 0.060 |
| **HC low - PD low** | 31.482 | -3.491 | **0.001** | **0.003** |
| **HC moderate - PD moderate** | 31.482 | -3.880 | **<0.001** | **0.001** |
| **HC high - PD high** | 31.482 | -5.713 | **<0.001** | **<0.001** |
| **P08** |  |  |  |  |
|  | **Num df** | **Den df** | **F value** | **p** |
| **Target location** | 1 | 47.387 | 3.593 | 0.064 |
| **Group** | 1 | 7.985 | 2.193 | 0.177 |
| **PriorZone** | 3 | 964.842 | 40.085 | **<0.001** |
| **Viewing** | 1 | 31.528 | 2.599 | 0.117 |
| **Target location:Group** | 1 | 967.726 | 13.677 | **<0.001** |
| **Target location:PriorZone** | 3 | 964.842 | 15.941 | **<0.001** |
| **Group:PriorZone** | 3 | 964.842 | 3.298 | **0.020** |
| **Target location:Group:PriorZone** | 3 | 964.842 | 3.285 | **0.020** |
| **Target location = expected** | | | |  |
|  | **Chi-square** | **Df** | **p** |  |
| **Group** | 6.749 | 1 | **0.009** |  |
| **PriorZone** | 206.477 | 3 | **<0.001** |  |
| **Viewing** | 0.050 | 1 | 0.823 |  |
| **Group:PriorZone** | 14.633 | 3 | **0.002** |  |
| **Target location = unexpected** | | | |  |
|  | **Chi-square** | **Df** | **p** |  |
| **Group** | 0.169 | 1 | 0.681 |  |
| **PriorZone** | 55.600 | 3 | **<0.001** |  |
| **Viewing** | 4.814 | 1 | **0.028** |  |
| **Group:PriorZone** | 4.372 | 3 | 0.224 |  |
| **Pairwise comparisons (Target location = expected)** | | | | |
| **contrast** | **df** | **t value** | **uncorrected p** | **Holm-corrected p** |
| **HC zero - PD zero** | 22.193 | -1.607 | 0.122 | 0.367 |
| **HC low - PD low** | 22.193 | -0.980 | 0.337 | 0.481 |
| **HC moderate - PD moderate** | 22.193 | -1.206 | 0.241 | 0.481 |
| **HC high - PD high** | 22.193 | -3.984 | **0.001** | **0.002** |
| **Pairwise comparisons (Target location = unexpected)** | | | | |
| **contrast** | **df** | **t value** | **uncorrected p** | **Holm-corrected p** |
| **HC zero - PD zero** | 17.772 | -1.098 | 0.287 | 1.000 |
| **HC low - PD low** | 17.772 | -0.439 | 0.666 | 1.000 |
| **HC moderate - PD moderate** | 17.772 | 0.468 | 0.645 | 1.000 |
| **HC high - PD high** | 17.772 | -0.223 | 0.826 | 1.000 |
| **P09** |  |  |  |  |
|  | **Num df** | **Den df** | **F value** | **p** |
| **Target location** | 1 | 52.823 | 0.392 | 0.534 |
| **Group** | 1 | 7.930 | 23.885 | **0.001** |
| **PriorZone** | 3 | 968.859 | 70.883 | **<0.001** |
| **Viewing** | 1 | 31.749 | 0.228 | 0.636 |
| **Target location:Group** | 1 | 970.883 | 1.448 | 0.229 |
| **Target location:PriorZone** | 3 | 968.859 | 17.955 | **<0.001** |
| **Group:PriorZone** | 3 | 968.859 | 21.663 | **<0.001** |
| **Target location:Group:PriorZone** | 3 | 968.859 | 6.335 | **<0.001** |
| **Target location = expected** | | | |  |
|  | **Chi-square** | **Df** | **p** |  |
| **Group** | 30.233 | 1 | **<0.001** |  |
| **PriorZone** | 134.478 | 3 | **<0.001** |  |
| **Viewing** | 1.336 | 1 | 0.248 |  |
| **Group:PriorZone** | 60.288 | 3 | **<0.001** |  |
| **Target location = unexpected** | | | |  |
|  | **Chi-square** | **Df** | **p** |  |
| **Group** | 16.191 | 1 | **<0.001** |  |
| **PriorZone** | 56.792 | 3 | **<0.001** |  |
| **Viewing** | 3.460 | 1 | 0.063 |  |
| **Group:PriorZone** | 12.125 | 3 | **0.007** |  |
| **Pairwise comparisons (Target location = expected)** | | | | |
| **contrast** | **df** | **t value** | **uncorrected p** | **Holm-corrected p** |
| **HC zero - PD zero** | 43.505 | -0.469 | 0.642 | 0.642 |
| **HC low - PD low** | 43.505 | -2.887 | **0.006** | **0.018** |
| **HC moderate - PD moderate** | 43.505 | -2.134 | **0.038** | 0.077 |
| **HC high - PD high** | 43.505 | -8.683 | **<0.001** | **<0.001** |
| **Pairwise comparisons (Target location = unexpected)** | | | | |
| **contrast** | **df** | **t value** | **uncorrected p** | **Holm-corrected p** |
| **HC zero - PD zero** | 23.168 | -1.287 | 0.211 | 0.211 |
| **HC low - PD low** | 23.168 | -2.842 | **0.009** | **0.018** |
| **HC moderate - PD moderate** | 23.168 | -3.358 | **0.003** | **0.008** |
| **HC high - PD high** | 23.168 | -4.386 | **<0.001** | **0.001** |
| **P10** |  |  |  |  |
|  | **Num df** | **Den df** | **F value** | **p** |
| **Target location** | 1 | 46.148 | 0.014 | 0.905 |
| **Group** | 1 | 7.950 | 0.829 | 0.389 |
| **PriorZone** | 3 | 968.733 | 51.650 | **<0.001** |
| **Viewing** | 1 | 31.384 | 3.109 | 0.088 |
| **Target location:Group** | 1 | 969.947 | 0.018 | 0.894 |
| **Target location:PriorZone** | 3 | 968.733 | 5.730 | **0.001** |
| **Group:PriorZone** | 3 | 968.733 | 3.478 | **0.016** |
| **Target location:Group:PriorZone** | 3 | 968.733 | 0.807 | 0.490 |
| **Target location = expected** | | | |  |
|  | **Chi-square** | **Df** | **p** |  |
| **Group** | 0.974 | 1 | 0.324 |  |
| **PriorZone** | 202.909 | 3 | **<0.001** |  |
| **Viewing** | 0.056 | 1 | 0.813 |  |
| **Group:PriorZone** | 4.716 | 3 | 0.194 |  |
| **Target location = unexpected** | | | |  |
|  | **Chi-square** | **Df** | **p** |  |
| **Group** | 0.633 | 1 | 0.426 |  |
| **PriorZone** | 82.213 | 3 | **0.000** |  |
| **Viewing** | 4.720 | 1 | **0.030** |  |
| **Group:PriorZone** | 8.309 | 3 | **0.040** |  |
| **Pairwise comparisons (Target location = expected)** | | | | |
| **contrast** | **df** | **t value** | **uncorrected p** | **Holm-corrected p** |
| **HC zero - PD zero** | 20.916 | -0.293 | 0.773 | 1.000 |
| **HC low - PD low** | 20.916 | -1.176 | 0.253 | 0.759 |
| **HC moderate - PD moderate** | 20.916 | 0.047 | 0.963 | 1.000 |
| **HC high - PD high** | 20.916 | -1.565 | 0.133 | 0.530 |
| **Pairwise comparisons (Target location = unexpected)** | | | | |
| **contrast** | **df** | **t value** | **uncorrected p** | **Holm-corrected p** |
| **HC zero - PD zero** | 16.917 | -0.240 | 0.813 | 1.000 |
| **HC low - PD low** | 16.917 | -0.060 | 0.953 | 1.000 |
| **HC moderate - PD moderate** | 16.917 | -0.298 | 0.770 | 1.000 |
| **HC high - PD high** | 16.917 | -1.923 | 0.071 | 0.286 |
| **P11** |  |  |  |  |
|  | **Num df** | **Den df** | **F value** | **p** |
| **Target location** | 1 | 46.613 | 0.088 | 0.768 |
| **Group** | 1 | 7.953 | 0.440 | 0.526 |
| **PriorZone** | 3 | 968.740 | 61.722 | **<0.001** |
| **Viewing** | 1 | 31.388 | 3.405 | 0.074 |
| **Target location:Group** | 1 | 969.982 | 0.029 | 0.864 |
| **Target location:PriorZone** | 3 | 968.740 | 7.662 | **<0.001** |
| **Group:PriorZone** | 3 | 968.740 | 5.698 | **0.001** |
| **Target location:Group:PriorZone** | 3 | 968.740 | 0.035 | 0.991 |
| **Target location = expected** | | | |  |
|  | **Chi-square** | **Df** | **p** |  |
| **Group** | 0.416 | 1 | 0.519 |  |
| **PriorZone** | 221.516 | 3 | **<0.001** |  |
| **Viewing** | 0.172 | 1 | 0.678 |  |
| **Group:PriorZone** | 9.369 | 3 | **0.025** |  |
| **Target location = unexpected** | | | |  |
|  | **Chi-square** | **Df** | **p** |  |
| **Group** | 0.415 | 1 | 0.519 |  |
| **PriorZone** | 83.287 | 3 | **<0.001** |  |
| **Viewing** | 4.545 | 1 | **0.033** |  |
| **Group:PriorZone** | 7.817 | 3 | **0.050** |  |
| **Pairwise comparisons (Target location = expected)** | | | | |
| **contrast** | **df** | **t value** | **uncorrected p** | **Holm-corrected p** |
| **HC zero - PD zero** | 20.744 | 0.238 | 0.814 | 1.000 |
| **HC low - PD low** | 20.744 | -0.051 | 0.960 | 1.000 |
| **HC moderate - PD moderate** | 20.744 | -0.088 | 0.931 | 1.000 |
| **HC high - PD high** | 20.744 | -2.054 | 0.053 | 0.211 |
| **Pairwise comparisons (Target location = unexpected)** | | | | |
| **contrast** | **df** | **t value** | **uncorrected p** | **Holm-corrected p** |
| **HC zero - PD zero** | 17.017 | 0.156 | 0.878 | 1.000 |
| **HC low - PD low** | 17.017 | -0.277 | 0.785 | 1.000 |
| **HC moderate - PD moderate** | 17.017 | -0.174 | 0.864 | 1.000 |
| **HC high - PD high** | 17.017 | -1.744 | 0.099 | 0.397 |
| **P12** |  |  |  |  |
|  | **Num df** | **Den df** | **F value** | **p** |
| **Target location** | 1 | 44.736 | 0.716 | 0.402 |
| **Group** | 1 | 7.946 | 0.067 | 0.802 |
| **PriorZone** | 3 | 968.403 | 44.164 | **<0.001** |
| **Viewing** | 1 | 31.061 | 2.102 | 0.157 |
| **Target location:Group** | 1 | 969.956 | 1.471 | 0.225 |
| **Target location:PriorZone** | 3 | 968.403 | 8.960 | **<0.001** |
| **Group:PriorZone** | 3 | 968.403 | 1.409 | 0.239 |
| **Target location:Group:PriorZone** | 3 | 968.403 | 0.459 | 0.711 |
| **Target location = expected** | | | |  |
|  | **Chi-square** | **Df** | **p** |  |
| **Group** | 0.004 | 1 | 0.953 |  |
| **PriorZone** | 204.941 | 3 | **<0.001** |  |
| **Viewing** | 0.001 | 1 | 0.972 |  |
| **Group:PriorZone** | 1.186 | 3 | 0.756 |  |
| **Target location = unexpected** | | | |  |
|  | **Chi-square** | **Df** | **p** |  |
| **Group** | 0.283 | 1 | 0.595 |  |
| **PriorZone** | 82.120 | 3 | **<0.001** |  |
| **Viewing** | 3.636 | 1 | 0.057 |  |
| **Group:PriorZone** | 4.508 | 3 | 0.212 |  |
| **Pairwise comparisons (Target location = expected)** | | | | |
| **contrast** | **df** | **t value** | **uncorrected p** | **Holm-corrected p** |
| **HC zero - PD zero** | 20.582 | 0.295 | 0.771 | 1.000 |
| **HC low - PD low** | 20.582 | 0.292 | 0.773 | 1.000 |
| **HC moderate - PD moderate** | 20.582 | 0.085 | 0.933 | 1.000 |
| **HC high - PD high** | 20.582 | -0.492 | 0.628 | 1.000 |
| **Pairwise comparisons (Target location = unexpected)** | | | | |
| **contrast** | **df** | **t value** | **uncorrected p** | **Holm-corrected p** |
| **HC zero - PD zero** | 16.713 | -0.003 | 0.997 | 1.000 |
| **HC low - PD low** | 16.713 | 0.228 | 0.822 | 1.000 |
| **HC moderate - PD moderate** | 16.713 | -1.116 | 0.280 | 1.000 |
| **HC high - PD high** | 16.713 | -0.798 | 0.436 | 1.000 |
| **P13** |  |  |  |  |
|  | **Num df** | **Den df** | **F value** | **p** |
| **Target location** | 1 | 47.968 | 0.049 | 0.826 |
| **Group** | 1 | 7.999 | 1.027 | 0.341 |
| **PriorZone** | 3 | 964.704 | 24.009 | **<0.001** |
| **Viewing** | 1 | 31.305 | 2.981 | 0.094 |
| **Target location:Group** | 1 | 967.534 | 0.000 | 0.985 |
| **Target location:PriorZone** | 3 | 964.704 | 6.987 | **<0.001** |
| **Group:PriorZone** | 3 | 964.704 | 0.783 | 0.504 |
| **Target location:Group:PriorZone** | 3 | 964.704 | 0.037 | 0.991 |
| **Target location = expected** | | | |  |
|  | **Chi-square** | **Df** | **p** |  |
| **Group** | 1.136 | 1 | 0.286 |  |
| **PriorZone** | 198.410 | 3 | **<0.001** |  |
| **Viewing** | 0.127 | 1 | 0.721 |  |
| **Group:PriorZone** | 1.165 | 3 | 0.761 |  |
| **Target location = unexpected** | | | |  |
|  | **Chi-square** | **Df** | **p** |  |
| **Group** | 0.811 | 1 | 0.368 |  |
| **PriorZone** | 69.752 | 3 | **<0.001** |  |
| **Viewing** | 3.869 | 1 | **0.049** |  |
| **Group:PriorZone** | 1.302 | 3 | 0.729 |  |
| **Pairwise comparisons (Target location = expected)** | | | | |
| **contrast** | **df** | **t value** | **uncorrected p** | **Holm-corrected p** |
| **HC zero - PD zero** | 19.815 | 0.426 | 0.675 | 1.000 |
| **HC low - PD low** | 19.815 | 0.706 | 0.489 | 1.000 |
| **HC moderate - PD moderate** | 19.815 | 0.849 | 0.406 | 1.000 |
| **HC high - PD high** | 19.815 | 1.282 | 0.215 | 0.858 |
| **Pairwise comparisons (Target location = unexpected)** | | | | |
| **contrast** | **df** | **t value** | **uncorrected p** | **Holm-corrected p** |
| **HC zero - PD zero** | 16.948 | 0.258 | 0.800 | 1.000 |
| **HC low - PD low** | 16.948 | 0.821 | 0.423 | 1.000 |
| **HC moderate - PD moderate** | 16.948 | 0.708 | 0.489 | 1.000 |
| **HC high - PD high** | 16.948 | 1.072 | 0.299 | 1.000 |

Results of the statistical tests used to arrive the results in the section "Visual Search: Analyzing Individual Fixations in Prior Zones". Each patient’s normalized dwell time on prior zones (between start of the trial and response) is compared to the HC group.

p values lower than 0.05 are printed in bold.

Df/df: degrees of freedom
