## Supplementary figures and images for "Navigating Visual Challenges: How Parkinson’s Disease Alters Cognitive Priorities in Visual Search"

### Fig s1

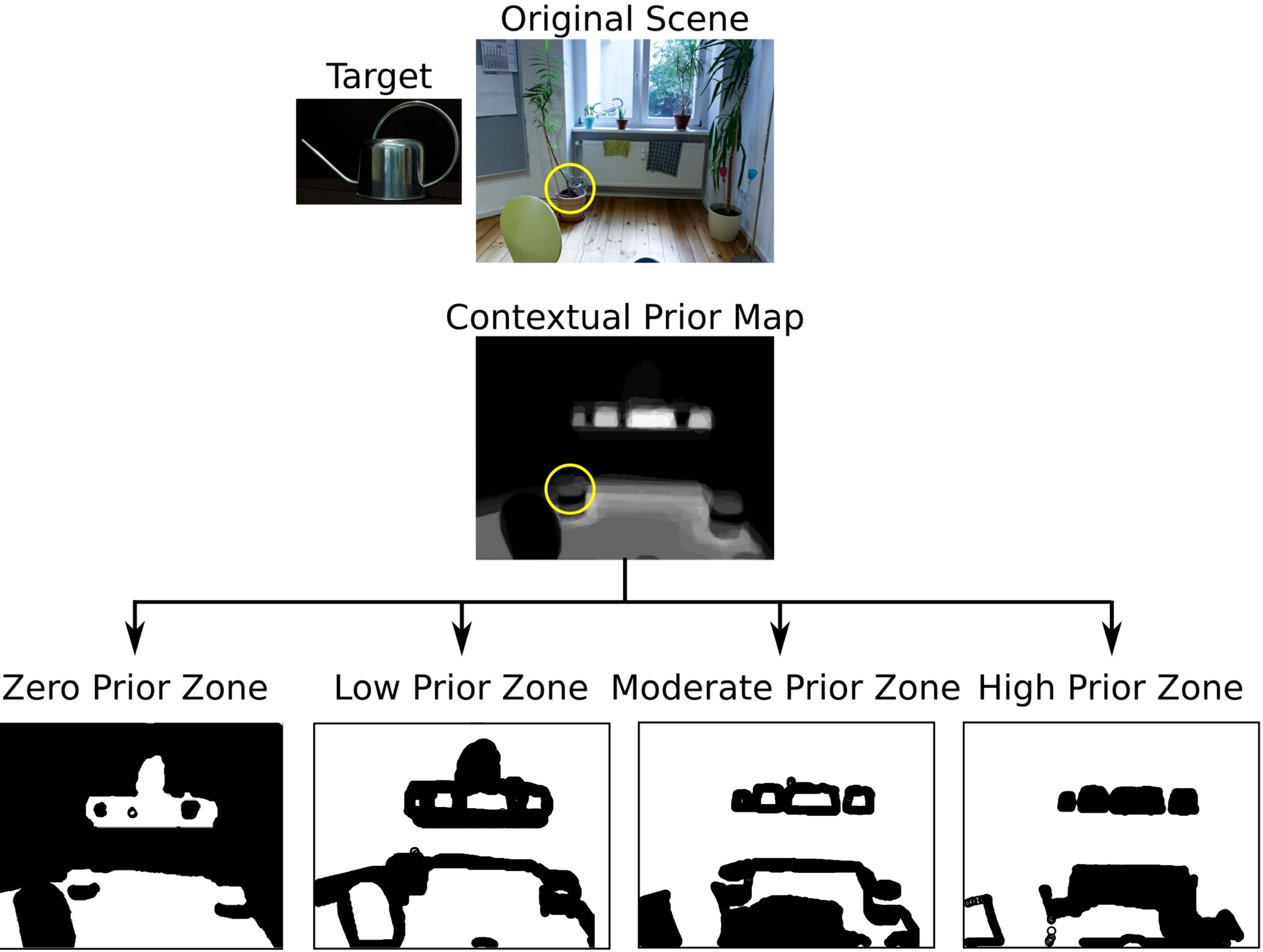
